# Per- and polyfluoroalkyl substances concentrations are associated with an unfavorable cardio-metabolic risk profile: findings from two population-based cohort studies

**DOI:** 10.1101/2023.10.19.23296512

**Authors:** Tariq O. Faquih, Elvire N. Landstra, Astrid van Hylckama Vlieg, N.Ahmad Aziz, Ruifang Li-Gao, Renée de Mutsert, Frits R. Rosendaal, Raymond Noordam, Diana van Heemst, Dennis O. Mook-Kanamori, Ko Willems van Dijk, Monique M.B. Breteler

## Abstract

Per- and polyfluoroalkyl substances (PFAS) are widely used and persistent chemicals, leading to ubiquitous exposure. Although high PFAS levels have been associated with an adverse cardiovascular risk profile, the distribution of levels and relations with cardio-metabolic risk markers in the general population have not been fully characterized. We assessed the association between blood levels of perfluorooctaneic acid (PFOA), perfluorooctane sulfonic acid (PFOS), and perfluorohexanesulfonic acid (PFHxS) and a range of lipoproteins and metabolites as well as clinical lipid measurements. We used data from participants of the Netherlands Epidemiology of Obesity study (NEO) (n= 584) and the Rhineland Study (n= 1,962), jointly spanning an age range of 30 to 89 years. PFAS were measured with the Metabolon HD4 platform, and lipoprotein and metabolite profiles were measured using Nightingale’s nuclear magnetic resonance-spectroscopy platform, and mainly comprised lipoprotein markers. Using linear regression analyses, we quantified age-, sex- and education-adjusted associations of PFOA, PFOS, and PFHxS with clinical lipid measurements and 224 lipoproteins and metabolites.

Higher levels of PFAS, particularly PFOS and PFHxS, were associated with higher concentrations of total lipid, cholesterol and phospholipid content in most HDL, IDL, LDL and VLDL subclasses. The effect sizes were age-dependent for the majority of the associations, with the deleterious effects of PFAS being generally stronger in people below compared to those above median age. Our observation that in the general population even low PFAS concentrations are associated with an unfavorable lipid profile, calls for further critical regulation of PFAS substances.

## 1. Introduction

Per- and polyfluoroalkyl substances (PFAS) are man-made chemicals that have been widely used in many industrial processes and products since the 1950s (Roth et al. 2021). These chemicals are persistent and resilient in nature, allowing them to circulate in water sources and become widespread around the globe, making them colloquially known as the ‘forever chemicals’ (RIVM ; Nordby, Luck 1956; Stockholm Convention on Persistent Organic Pollutants (POPs) 2001; Buck et al. 2011; Priestly 2018; Sunderland et al. 2019; Grandjean et al. 2020; Schrenk et al. 2020). High environmental PFAS levels may be caused by contamination in the vicinity of PFAS-producing factories, but also by the use and breakdown of PFAS-containing products—such as fire extinguishers, non-stick cooking pans, certain food packaging, textiles, clothing, cosmetics, and pesticides (Sunderland et al. 2019; Barhoumi et al. 2022). Human exposure can occur through contact with a contaminated environment (e.g. through dermal exposure or inhalation), contact with PFAS-containing products, as well as ingestion with drinking water and food (Sunderland et al. 2019). Given the persistence of PFAS, human exposure is ubiquitous and chronic. PFAS accumulate in the human body over time as they are broken down slowly, where the half-life of common PFAS is estimated to be between 4 and 8.5 years (Olsen et al. 2007).

Ever since the introduction of PFAS production, health concerns were raised (Nordby, Luck 1956; 3M Company Chemical Division 1963; Sherman 1973; Sunderland et al. 2019). The main focus has long been on direct exposure to high levels of PFAS, which was found to be associated with a range of adverse health outcomes, including obesity, kidney disease, cancer, thyroid disease, hypercholesteremia, dyslipidemia, liver damage, reduced antibody response to vaccination, and a higher risk of severe course of COVID-19 (Stockholm Convention on Persistent Organic Pollutants (POPs) 2001; Priestly 2018; Grandjean et al. 2020; Schrenk et al. 2020). Animal studies and studies in high-risk groups, such as children, who are at high risk of adverse effects, and high exposure groups, such as factory workers, have found PFAS to be related with changes in the immune system, proteome, hormones, and metabolome (Liu et al. 2022; Shih et al. 2022). However, evidence of the effects of PFAS in humans remains limited and more research about the presence and the effects of low exposure in the general population is warranted (Sunderland et al. 2019; European Food Safety Authority 2020).

The health concerns from PFAS exposure resulted in the classification of certain PFAS as persistent pollutants that require regulation. Specifically, the use of perfluorooctane sulfonic acid (PFOS) and perfluorooctanoic acid (PFOA) has been subjected to growing restrictions (Stockholm Convention on Persistent Organic Pollutants (POPs) 2001; Stockholm Convention on Persistent Organic Pollutants (POPs) 2017). However, despite the increasing scrutiny, many PFAS species remain unregulated and even PFOA and PFOS are not yet fully banned in the European Union (RIVM 2022a). Thus, and worsened by their long half-life, PFAS exposure remains a public health issue (Duffek et al. 2020; Schrenk et al. 2020; RIVM 2021).

Specific mechanisms through which PFAS exert their effects remain largely unclear. Previous studies have shown that PFAS are most consistently associated with changes in lipid metabolism, particularly higher cholesterol levels (Priestly 2018). Recent advances in high throughput metabolomic approaches allow the analysis of large lipoprotein and metabolite panels and enable more in-depth examinations of lipid species and lipoprotein subclasses. Indeed, a recent study investigated the effect of PFAS on an extensive lipid panel in a sample of 50-year old adults (Haug et al. 2023). We aim to corroborate and expand upon that work by including a larger group and a wider age range. Especially, we aim to evaluate PFAS exposure levels in the general population and investigate their association with metabolites and lipoproteins, using clinical lipid measurements and targeted metabolomics. To enhance generalizability and robustness of the results, we used two different study populations: the Netherlands Epidemiology of Obesity study (NEO) (n = 584) and the Rhineland Study (n = 1,962).

## 2. Methods

### 2.1. Study Populations

#### 2.1.1. Netherlands Epidemiology of Obesity Study

NEO is a population-based, prospective cohort study of individuals aged 45–65 years, with an oversampling of individuals who are overweight or have obesity. Men and women aged between 45 and 65 years with a self-reported body mass index (BMI) of 27 kg/m^2^ or higher, living in the greater area of Leiden (in the West of the Netherlands) were eligible to participate in the NEO study. In addition, all inhabitants aged between 45 and 65 years from one municipality (Leiderdorp) were invited, irrespective of their BMI. Recruitment of participants started in September 2008 and was completed at the end of September 2012. In total, 6,671 participants have been included. Participants were invited to come to the NEO study center of the LUMC for one baseline study visit, where a blood sample of 108 mL was taken from the participants after an overnight fast of at least 10 hours (de Mutsert et al. 2013). In the current study, the samples of a random subset of 599 participants from the Leiderdorp sub-population (n=1,671) were used to measure untargeted metabolomic data. Among these individuals, 4 did not have the study outcome (targeted metabolomics) data, 2 were excluded due to missing data on educational level, and 9 were excluded due to measurement errors, leading to a final sample size of n = 584 (**Figure 1**).

**Figure 1:**
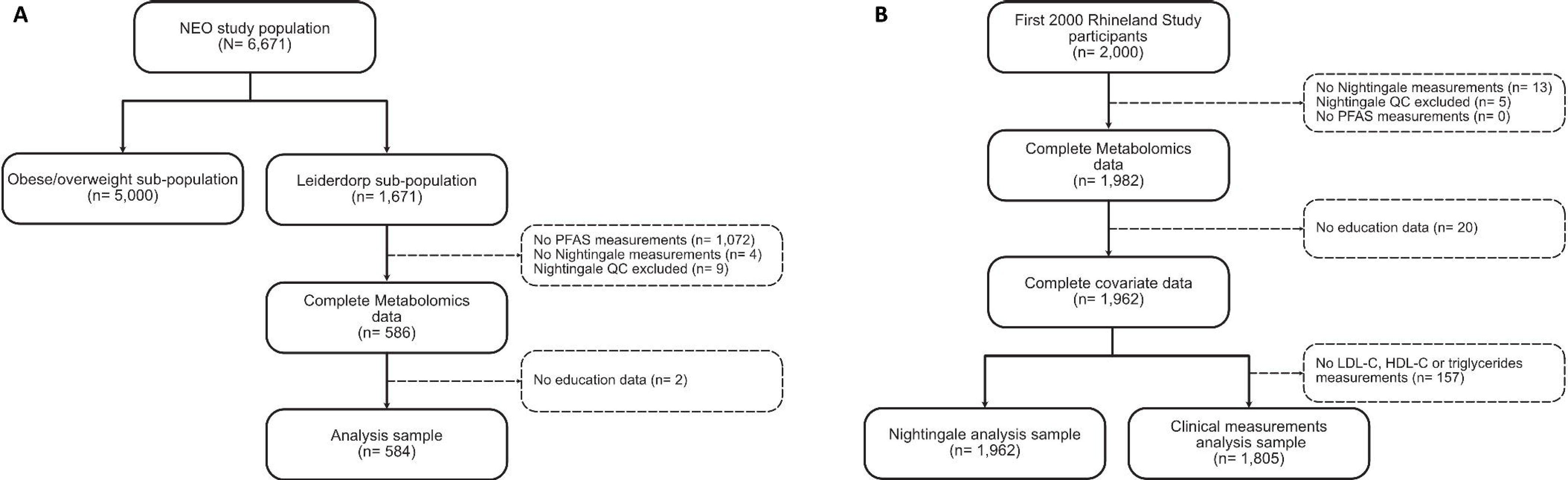
Overview of study population selection for NEO (A) and the Rhineland Study (B)

#### 2.1.2. Rhineland Study

The Rhineland Study is an ongoing community-based cohort study located in two geographically defined areas in Bonn, Germany. Participation is possible by invitation only. To be able to participate, participants had to be above 30 years of age at baseline and have sufficient command of the German language to provide written informed consent. Nightingale measurements were performed in plasma from the first 2,000 consecutive participants, 1,982 of which passed quality control. We further excluded 20 participants due to missing data on educational level, leaving a sample size of n=1,962 to be included in the analyses (**Figure 1**). Of these, 1,805 participants had complete data on LDL, HDL, total triglyceride, and cholesterol levels, which were used for the analysis of clinical lipid measurements.

### 2.2. PFAS measurements

Blood PFAS concentrations were measured using the untargeted Metabolon™ Discovery HD4 platform at Metabolon Inc. (Durham, North Carolina, USA). In brief, this process involves four independent ultra-high-performance liquid chromatography mass spectrometry (UHPLC-MS/MS) platforms (Evans et al. 2014; Rhee et al. 2019). It uses two positive ionization reverse phase chromatography, one negative ionization reverse phase chromatography, and one hydrophilic interaction liquid chromatography negative ionization (Rhee et al. 2019). In NEO, PFOA and PFOS levels were quantified from fasting state serum samples, while in the Rhineland Study PFOA, PFOS and perfluorohexanesulfonic acid (PFHxS) concentrations were obtained from fasting plasma samples. PFAS were quantified as relative concentrations using the mass and charge peaks of the molecules. PFHxS was not measured in NEO, because it was not detectable in serum samples after the necessary sample preparations. To ensure comparability across studies, we Z-standardized all PFAS levels before subsequent analyses. The distribution of PFAS levels in the different studies can be found in Supplementary Figure 1.

### 2.3. Metabolic and lipoprotein measurements

Clinical lipids, i.e. total serum levels of LDL, HDL and total cholesterol, and triglycerides, were measured in NEO as described previously (de Mutsert et al. 2013). In the Rhineland Study, serum levels of LDL, HDL and total cholesterol, and triglycerides, were measured using routine methods at the University Hospital in Bonn.

Fasting serum (NEO) and plasma (Rhineland Study) metabolite levels were quantified using the Nightingale nuclear magnetic resonance untargeted metabolomic platform (Nightingale Health Ltd, Helsinki, Finland), which quantifies 224 metabolites and metabolite ratios. The platform predominantly provides detailed lipoprotein lipid information, in addition to absolute concentrations of various other metabolites, including amino acids, free fatty acids and ketone bodies (Soininen et al. 2015).

### 2.4. Assessment of covariates

In both the Rhineland Study and NEO, questionnaire and food frequency questionnaires were used to collect demographic and lifestyle information, including current smoking status (yes/no), alcohol intake (g/day), and education (low/middle/high). In the Rhineland Study, missing values (n= 163) for smoking were imputed based on HD4-measured cotinine levels in the blood according to the method described by St Helen et al. (St Helen et al. 2012). To assure the quality of the data, alcohol values belonging to participants reporting an overall improbable caloric intake (<600 or >8,000) were excluded, as per the method of Galbete et al. (Galbete et al. 2018). The name, dosage, anatomical therapeutic chemical (ATC) code and prescription status of each medication that participants were currently taking, or had taken in the past year, were recorded in an interview. Use of lipid-lowering mediation was based on ATC code C10 in the Rhineland Study and ATC codes C10AA or C10BA in NEO.

The International Standard Classification of Education 2011 (ISCED) (UNESCO Institute for Statistics 2012) was used to standardize education across both studies. Participants’ education level was reclassified as low (lower secondary education or below), middle (upper secondary education to undergraduate university level) or high (postgraduate university study) in both studies.

### 2.5. Statistical Analysis

#### 2.5.1. Imputation of Missing Metabolite Values

A total of 224 metabolites were measured on the targeted metabolomics Nightingale platform. These metabolites had missing values ranging from 0.3% to 9%. Missing values were set to 0 where the levels were considered to be below detection. Missingness due to other reasons (according to the flagging of missing status by Nightingale, e.g. as ‘equipment failure’) was imputed using a previously described pipeline (Faquih et al. 2020). Accordingly, imputed datasets were generated using multiple imputation. To ensure normality, all metabolites were log-transformed after adding 1 to account for 0s.

#### 2.5.2. Linear Regression Models

Multiple linear regression models were used to associate the log-transformed and Z-standardized PFAS concentrations (exposures) with the log-transformed Nightingale and clinical lipid measurements (outcomes). In our first models, we adjusted for biological sex (women/men), age (years) and education (low/middle/high). As PFAS accumulate during the lifespan (Pérez et al. 2013) and may have different effects across age groups, we next included a multiplicative interaction term between age and the PFAS substances. We also assessed possible effect differences between men and women (Koskela et al. 2022) by additionally including a sex-interaction term to our models. If the interaction estimate was significant, we performed a stratified analysis on the basis of sex or the median age (54 years). For ease of comparability in the figures, analyses were run on the Z-standardized metabolite levels after the log transformation. To test whether the associations were dependent on traditional risk behaviors and medication use, we ran additional analyses adjusting for alcohol intake, smoking, BMI and lipid-lowering medication. Moreover, we assessed the robustness of our results by performing a sensitivity analysis where we truncated extreme outlier values (>5 standard deviations (SDs)) in the PFAS levels.

#### 2.5.3. Multiple testing correction

As the Nightingale measurements are inherently highly correlated, we used the method described by Li and Ji (Li, Ji 2005) to calculate the effective number of independent variables. Accordingly, the number of independent Nightingale variables was estimated at 66. We furthermore included 4 clinical lipid measurements as outcomes. Thus, we considered a p-value < 0.0007 (0.05/70) as statistically significant.

All analyses were performed using R (R Core Team 2019) v4.1.0 (2021-05-18) in NEO and v4.0.5 (2021-03-31) in the Rhineland Study. Figures were created using the *ggplot2* (Hadley 2016) R package.

## 3. Results

### 3.1. Population characteristics

Participants in the Rhineland Study (57% women) had a median age of 54 years (range: 30 – 89), an average BMI of 25.8 (SD: 4.5), and a relatively high education level (**Table 1**). NEO participants (52.6% women) had a median age of 54 years (range: 45 – 66), an average BMI of 25.9 (SD: 4.0), and were mostly of a medium education level. Average alcohol consumption in the Rhineland Study was higher than in the NEO study. Current smokers made up 13.7% of the Rhineland Study and 11.3% of NEO. PFOA and PFOS were measurable in all 599 NEO participants. In the Rhineland Study, PFOA, PFOS, and PFHxS were below the level of detection in none, 17, and three out of 1,962 participants, respectively.

**Table 1:**
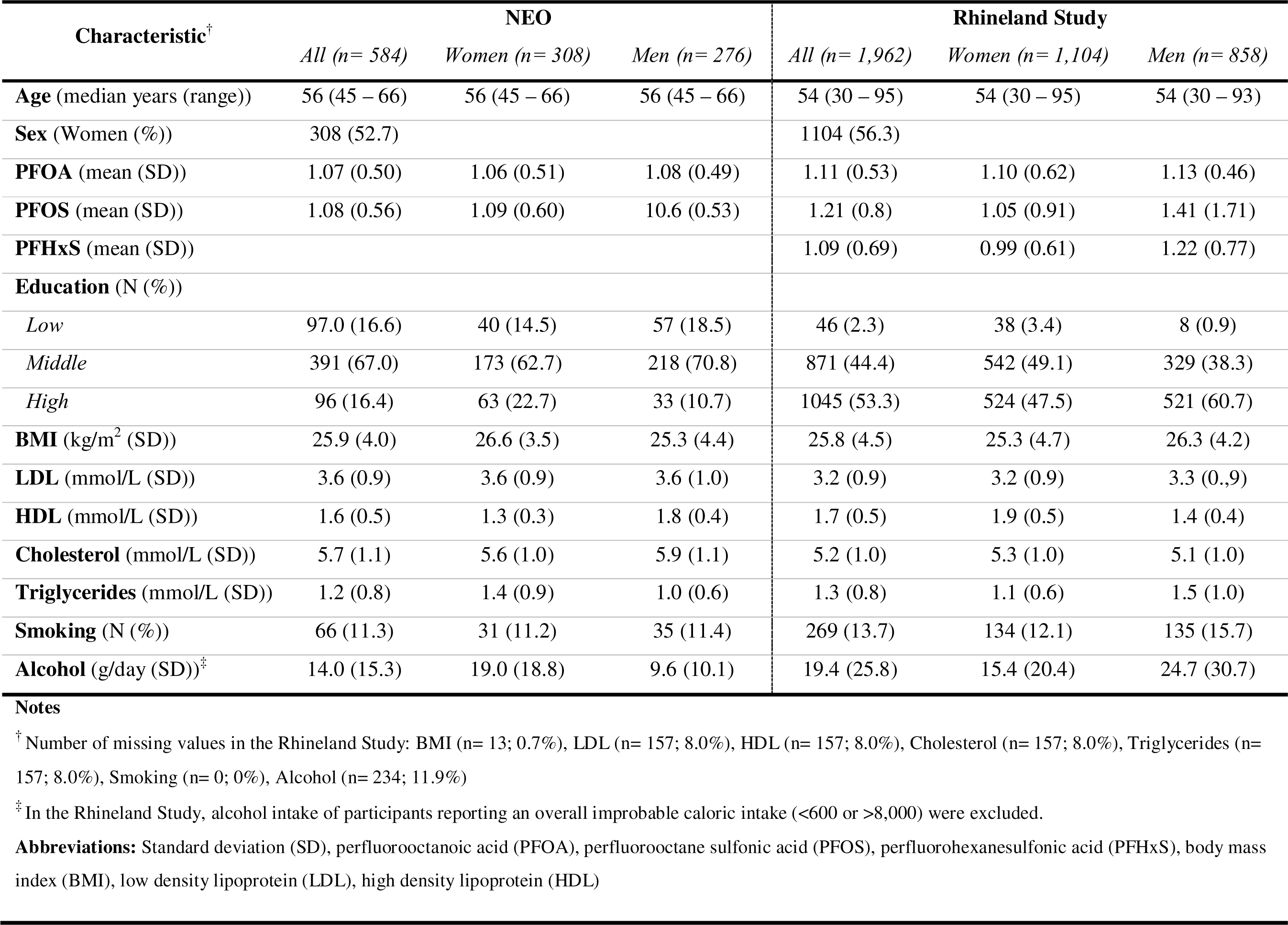
Population characteristics of NEO and the Rhineland Study.

### 3.2. PFAS and clinical lipids

In the overall analyses, no associations were found between PFOA, PFOS or PFHxS and the clinical lipid measurements when adjusting for age, sex and education (**Table 2**). While this did not differ between men and women, we found statistically significant age-interaction effects for some of the associations (Rhineland Study: PFOS and PFHxS with total cholesterol, PFOA and PFOS with LDL-C; NEO: PFOS with total cholesterol and LDL-C). In the subsequent age-stratified analyses (≤54 years; >54 years) we only found an association of PFOS with higher cholesterol levels in the younger age group in NEO (**Supplementary Table 1**, **Supplementary Table 2**).

**Table 2:** Relation of blood PFAS levels with clinical lipid measurements (log-transformed) in NEO and the Rhineland Study.

| PFAS | Study | Model<sup>†</sup> | LDL-C |  | HDL-C |  | Total cholesterol |  | Triglycerides |  |
| --- | --- | --- | --- | --- | --- | --- | --- | --- | --- | --- |
| | | | $\beta$ [95%CI] | *p*-value<sup>‡</sup> | $\beta$ [95%CI] | *p*-value<sup>‡</sup> | $\beta$ [95%CI] | *p*-value<sup>‡</sup> | $\beta$ [95%CI] | *p*-value<sup>‡</sup> |
| PFOA | NEO | Overall | 0.019<br>[ 0.001; 0.036] | 0.0413 | 0.016<br>[ 0.004; 0.029] | 0.0112 | 0.020<br>[ 0.007; 0.034] | 0.0025 | 0.013<br>[-0.010; 0.036] | 0.2787 |
|  | Rhineland Study | Overall | 0.005<br>[-0.006; 0.016] | 0.388 | 0.007<br>[-0.001; 0.016] | 0.0690 | 0.008<br>[-0.001; 0.016] | 0.0665 | -0.001<br>[-0.015; 0.014] | 0.9362 |
| PFOS | NEO | Overall | 0.023<br>[ 0.004; 0.041] | 0.0153 | 0.007<br>[-0.006; 0.021] | 0.2675 | 0.020<br>[ 0.007; 0.034] | 0.0037 | 0.016<br>[-0.009; 0.040] | 0.2031 |
|  | Rhineland Study | Overall | 0.007<br>[-0.003; 0.017] | 0.1726 | 0.005<br>[-0.002; 0.013] | 0.1575 | 0.007<br>[-0.001; 0.014] | 0.0837 | -0.005<br>[-0.018; 0.008] | 0.4502 |
| PFHxS | Rhineland Study | Overall | 0.013<br>[0.003; 0.023] | 0.0128 | 0.012<br>[0.004; 0.019] | 0.0024 | 0.012<br>[0.004; 0.020] | 0.0023 | -0.006<br>[-0.019; 0.007] | 0.3969 |

**Notes**
<sup>‡</sup>A *p* value of < 0.0007 was considered significant after accounting for multiple testing using the Li and Ji method
<sup>†</sup> Models were sex-, age- and education-adjusted.
**Abbreviations:** Beta estimate ( $\beta$ ), confidence interval (CI), perfluorooctanoic acid (PFOA), perfluorooctane sulfonic acid (PFOS), perfluorohexanesulfonic acid (PFHxS), low density lipoprotein (LDL), high density lipoprotein (HDL)

### 3.3. PFAS and metabolic profiles

Overall, all PFAS substances were usually, albeit largely non-significantly, associated with higher levels of different cholesterols, fatty acids and the lipid content of differently-sized IDL, LDL and VLDL subclasses after adjustment for sex, age and education. Statistically significant associations were observed between PFOA and a higher concentration and lipid content of small HDL particles in NEO and between PFOA and sphingomyelins, cholesterol in HDL3, and albumin in the Rhineland Study. PFOS was statistically significantly associated with higher fatty acid levels in NEO, while PFHxS was statistically significantly associated with higher levels of cholesterols, fatty acids, albumin, and apolipoprotein A1 (apoA1), and with lower levels of the amino acid phenylalanine in the Rhineland Study (**Figure 2**).

**Figure 2:**
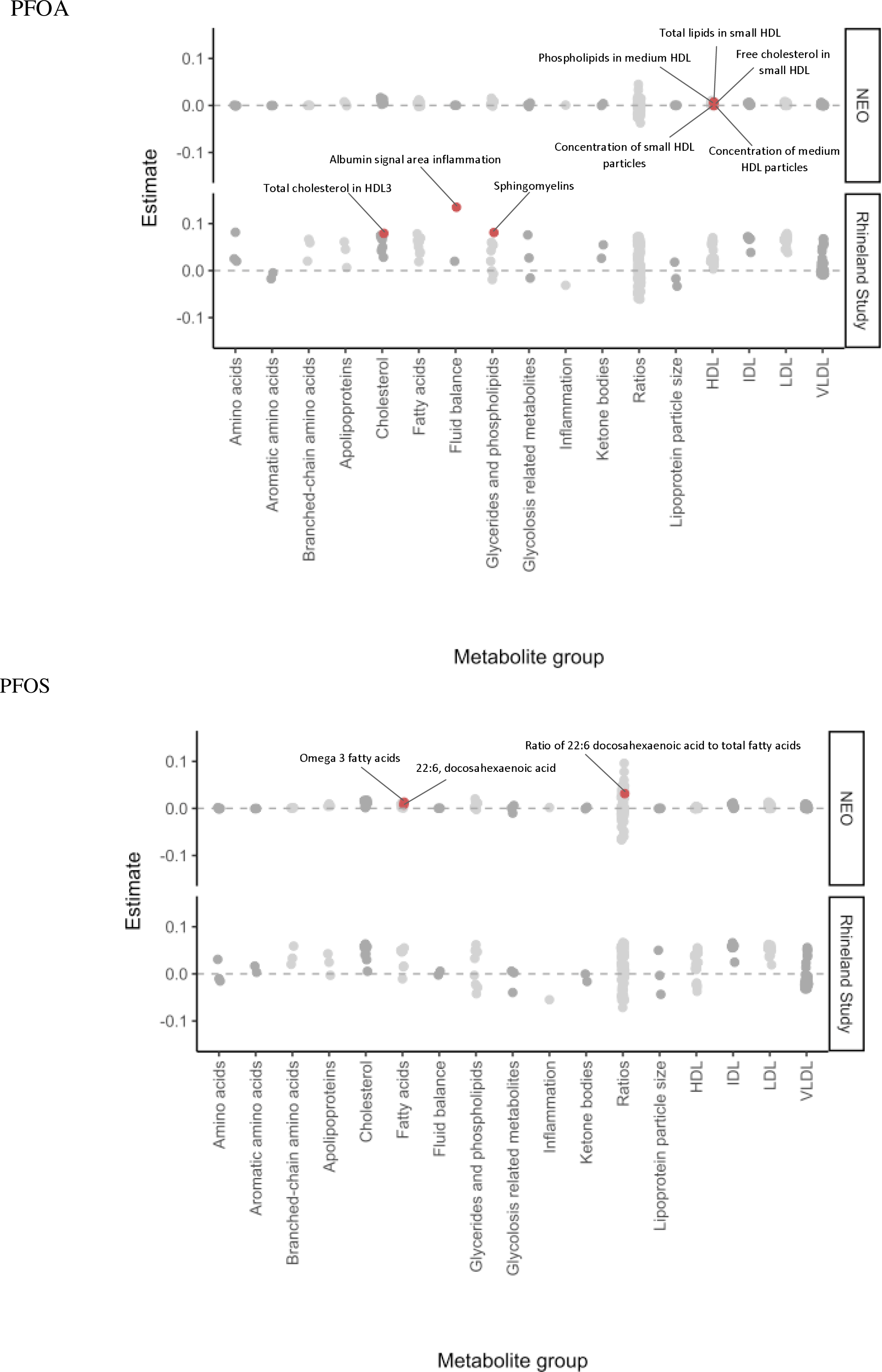
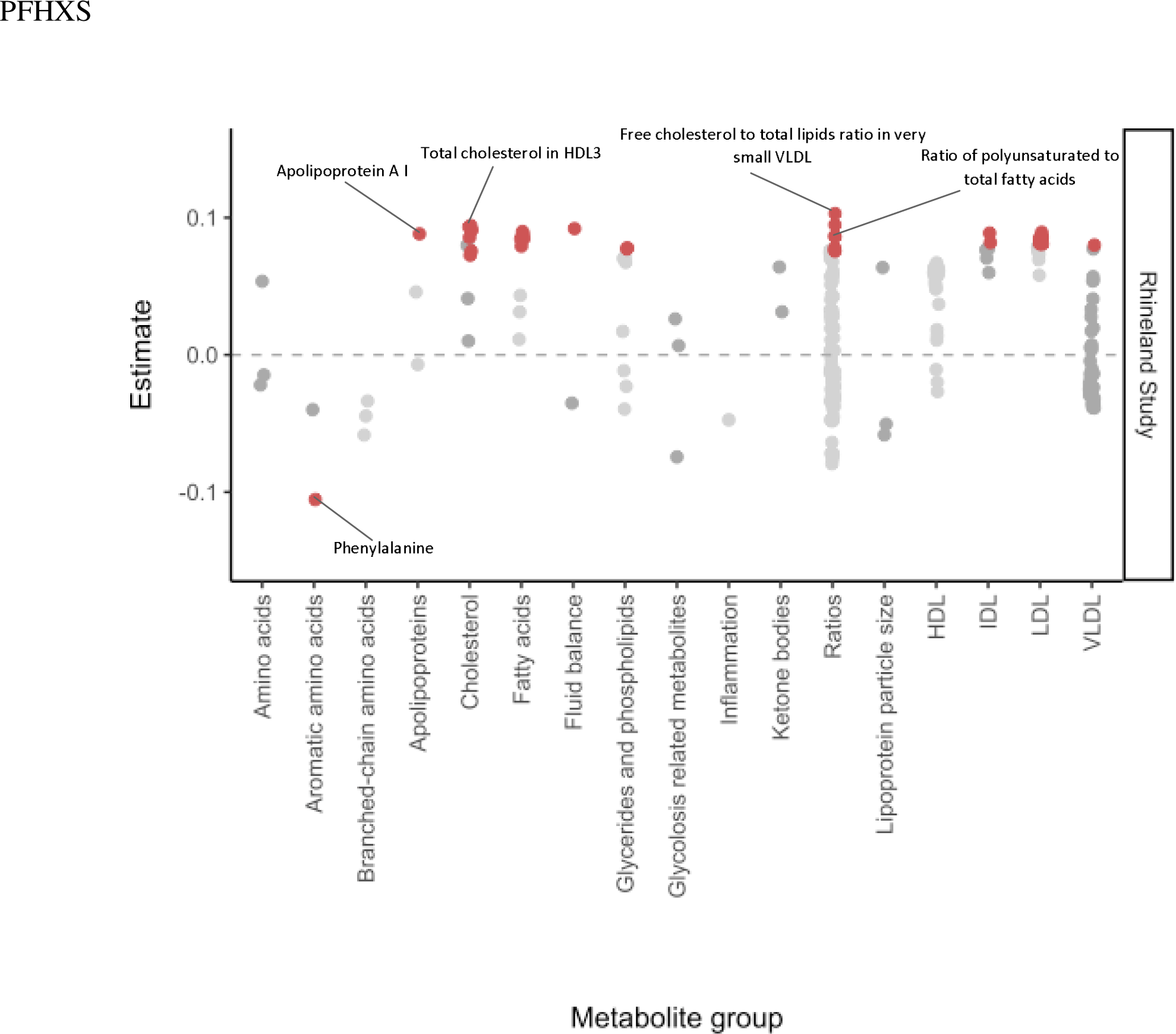
Plotted beta estimates and their distribution per metabolite class for NEO and the Rhineland Study for PFOA (A), PFOS (B), and PFHxS (C), where the top 5 significant hits are annotated

We found significant age-interactions for PFOA (NEO: 5; Rhineland Study: 1) and PFHxS (Rhineland Study: n= 8), and many for PFOS (NEO: 54; Rhineland Study: 80) in both studies. These age-effects were observed for almost all lipoprotein and metabolite classes, including cholesterol, cholesterol ester and total lipid content of IDLs, LDLs, and VLDLs, as well as apolipoprotein B (apoB), fatty acids, amino acids, albumin, and sphingomyelin levels. The associations were almost always stronger in the younger compared to the older age group (**Figure 3**).

**Figure 3:**
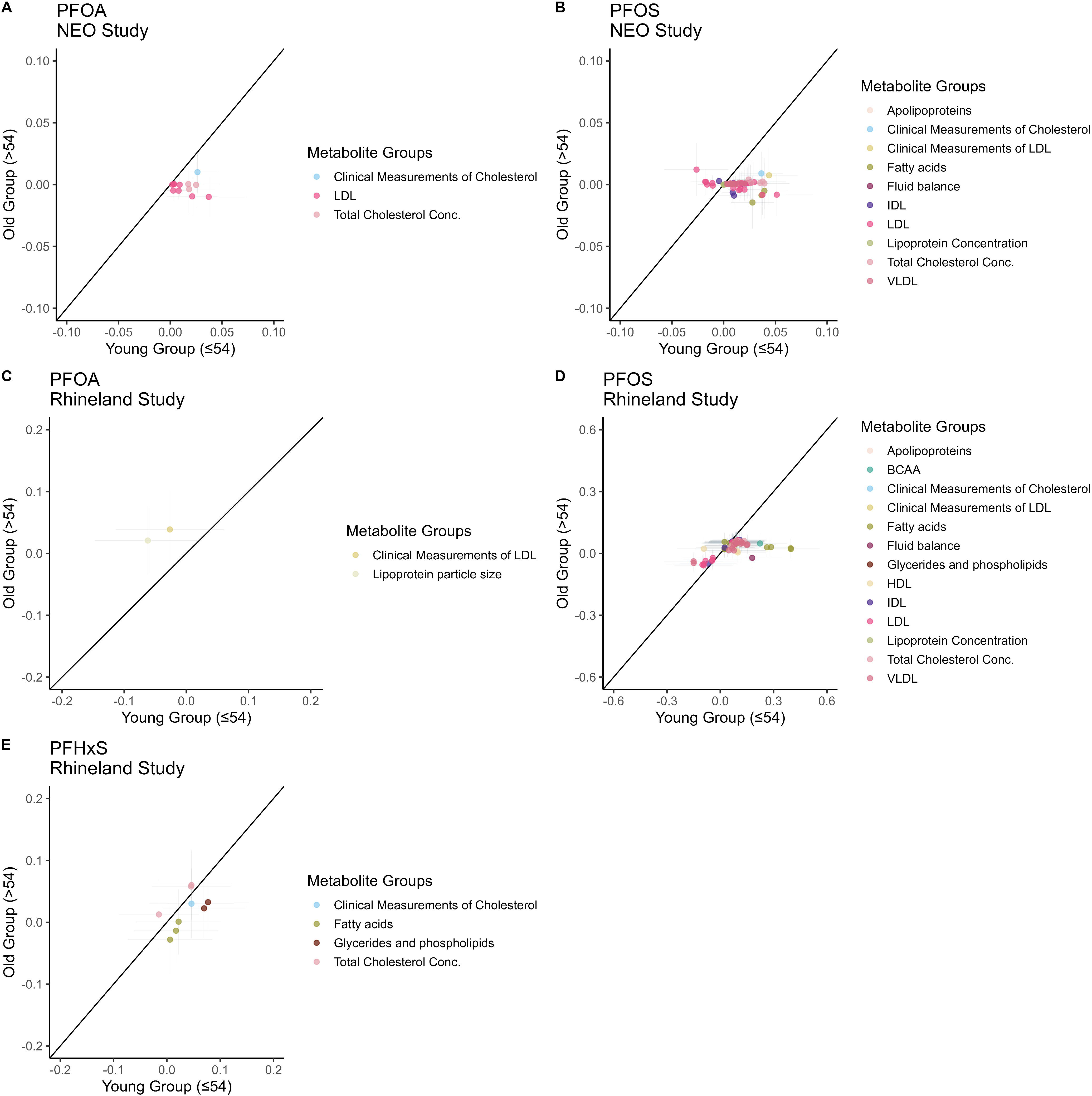
Beta/beta plots showing the beta estimates in the young (≤54 years) versus old (>54 years) group where the age interaction term was significant for NEO (A, B) and the Rhineland Study (C, D, E)

Lastly, we checked whether the above-mentioned associations were confounded by smoking, alcohol consumption, BMI, and lipid-lowering medication (**Supplementary Table 1**, **Supplementary Table 2**). In NEO, the associations remained largely unchanged. In the Rhineland Study, the number of significant associations changed across all PFAS substances. However, while additional adjustment decreased the strength of the effect, particularly when including BMI or alcohol intake, the previously described patterns remained largely the same.

Truncating outliers of PFAS levels did not substantially change the results in NEO, but slightly increased the number of significant associations in the Rhineland Study (**Supplementary Table 1**, **Supplementary Table 2**), indicating that the observed associations were not driven by outliers.

## 4. Discussion

### 4.1. Summary

In this study, we investigated the association of three PFAS substances with clinically measured lipid biomarkers and a wide range of metabolites (n= 224) in the general population. By combining findings from the NEO Study (n= 584) and the Rhineland Study (n= 1,962), we report common and clinically relevant effects of PFAS on measures of lipid metabolism. Specifically, PFOS and PFHxS were associated with an adverse metabolomic profile characterized by increased levels of apoB, phosphoglycerides, total lipids, fatty acids, and the lipid content of LDL, IDL, and VLDL particles. The effect estimates were consistent with regard to both direction and magnitude in both cohorts and were not driven by outliers, further supporting the validity of our findings. Thus, we interpret our data to indicate that even low PFAS levels in the general population can have a detrimental effect on lipid metabolism.

### 4.2. Widespread PFAS exposure in the Netherlands and Germany

PFAS exposure is widespread in both Germany (Brede et al. 2010; Duffek et al. 2020; German Environment Agency 2020; Kotthoff et al. 2020) and the Netherlands (Gebbink, van Leeuwen 2020; RIVM 2021; van der Aa 2021; RIVM 2022b). Despite the slow phasing out of the production of PFAS substance in the Netherlands (Municipality of Dordrecht 2022), both surface and ground water, soil, vegetation, fish, and stock animals in the area remain highly contaminated. In addition, contamination in surface and drinking water was detected across the western regions of the Netherlands (Gebbink, van Leeuwen 2020). Recently, the National Institute for the Public Health and the Environment (RIVM) concluded that the levels of PFAS in the Netherlands are highly concerning and require further research (RIVM 2021). PFAS exposure is also a problem in Germany, where areas along the Rhine among others are marked as high exposure locations (Brendel et al. 2018; Kotthoff et al. 2020). Importantly, PFAS levels are detectable in the groundwater of most and soil samples of all states (German Environment Agency 2020). Indeed, in line with the high contamination levels of both countries and the long half-life of PFAS in human serum of between 4 and 8.5 years (Olsen et al. 2007; Schrenk et al. 2020), we found that PFAS levels were detectable in the blood of nearly all included participants from NEO and the Rhineland Study.

### 4.3. PFAS levels associated with metabolomic profiles of increased risk of cardiovascular disease

We found that even low PFAS levels were associated with a distinctive lipid profile, characterized by higher cholesterol and lipid content in HDL, IDL, LDL, and VLDL subclasses. We also found associations between PFAS levels and apoB, fatty acids, phosphoglycerides, IDL, and phospholipids, which is largely in line with a recent, smaller study among 50-year-old individuals in Sweden (Haug et al. 2023). A higher lipid content of lipoproteins has been implicated in cardiovascular disease (CVD) (Xiao et al. 2016), while higher levels of fatty acids and apoB have been consistently associated with myocardial infarction (Julkunen et al. 2022). Other studies have linked a similar metabolomic profile to a higher risk of cardiometabolic diseases such as CVD (Tikkanen et al. 2021), hypertension (Palmu et al. 2022), type 2 diabetes (Ahola-Olli et al. 2019), and non-alcoholic fatty liver disease (Sliz et al. 2018). Therefore, our results suggest that PFAS exposure may increase the risk of cardiometabolic outcomes by impacting the lipoprotein profile. Of note, we found that PFHxS, one of the often-used more recent substitutes of PFOA and PFOS (Sunderland et al. 2019; Li et al. 2020), affected metabolite concentrations as well as lipoprotein composition and concentrations in the same way as PFOA and PFOS. Additional adjustment for certain risk behaviors (alcohol consumption, smoking), obesity and lipid-lowering medication, generally decreased the strength of and number of significant associations, but did not substantially change the overall patterns. Thus, the risks associated with PFAS are largely independent of these other factors.

### 4.4. Effects of PFAS are partially dependent on age

Our population studies jointly covered a wide age-range (30-89 years), which allowed us to investigate whether the effects of PFAS levels in adults differed across age. We found significant age-interactions for most of the PFOA and PFOS associations with lipoproteins, with the effects generally being larger in younger individuals. On the other hand, the associations of PFHxS with metabolite outcomes were less dependent on age. It is unclear what underlies the stronger effects of PFOA and PFOS in younger people. A possible explanation could be the presence of more competing causes in older people, including changes in cholesterol and other lipid levels later in life due to medication use, change in body composition, hormones or diet (Ettinger et al. 1992; Ferrara et al. 1997; Downer et al. 2014; Zhang et al. 2020). Alternatively, it is possible that the timing and duration of a person’s exposure impact the effect. For example, early life exposure reportedly has more severe consequences than late-life exposure (Canova et al. 2021). Finally, it may be that the intensity of exposure, rather than cumulative exposure, is an important determinant of the physiological impact of PFAS exposure. Due to the long half-life, the same blood exposure level may result from lower exposures over a longer time period in older compared to younger persons. This might also explain why no clear age-effects were seen for PFHxS: as that has been in use much shorter, blood levels may reflect more similar exposure patterns between younger and older persons.

### 4.5. Strengths and limitations

Our main limitation stems from the cross-sectional nature of our data. As such, we cannot establish a causal link between the PFAS exposures and the metabolites. Nonetheless, reverse causation is unlikely due to the nature of the associations, as it is highly unlikely that lipoprotein concentrations would affect PFAS levels. Other limitations include the use of different sample media for the measurement of PFAS and Nightingale metabolites in NEO versus the Rhineland Study, as well as the use of relative, rather than absolute, PFAS levels. A strength of our study is that we comprehensively evaluated the effect of PFAS exposure in relation to a detailed lipid profile and a large variety of metabolites across a wide age-range, which has not been assessed before. Moreover, we found consistent and robust associations of PFAS levels with a cardio-metabolic risk profile across two independent European studies, which supports the reliability of our findings. Moreover, our results are in line with findings from a smaller, recent study (Haug et al. 2023), which further corroborates the validity of our results.

### 4.6. Conclusion

In conclusion, we found that even low levels of PFAS are associated with a detrimental lipid profile in the general population. We report associations between PFAS and higher apoB and lipid content, particularly cholesterol content, of most subclasses of IDL, LDL, and VLDL. Our results corroborate and expand on previous findings by showing a clear link between PFAS levels and an adverse lipid profile across different study populations, even at low PFAS levels. In the Netherlands, pre-determined cut-offs for safe levels of PFAS were recently used to conclude that the PFAS levels in the Westerschelde were no cause for concern (RIVM 2022b). However, our results indicate that there may be no safe levels below which exposure is without health hazard. Moreover, we found that a more recently developed PFAS, PFHxS, was as detrimental for health as were older PFAS (PFOA and PFOS). Thus, stricter regulations may be required for all PFAS substances. Furthermore, due to the persistent nature of PFAS and their recirculation in the environment, there is a need to actively remove these chemicals from the environment—methods for which are under development (Trang et al. 2022). The combination of the well-documented persistence of PFAS and their harmful effects ensures that exposure to these substances is an enduring public health concern, unless and until we find ways to effectively eliminate PFAS from our environment.

## Acknowledgements

We would like to thank all participants and the study personnel of the Rhineland Study. The authors of the NEO study thank all participants, all participating general practitioners for inviting eligible participants, all research nurses for data collection, and the NEO study group: Pat van Beelen, Petra Noordijk, and Ingeborg de Jonge for study coordination, laboratory, and data management.

## Statements and Declarations

### Author Contributions

**T.O. Faquih:** Conceptualization, Methodology, Formal Analysis, Writing – Original Draft Preparation, Visualization; **E.N. Landstra.:** Conceptualization, Methodology, Formal Analysis, Writing – Original Draft Preparation, Visualization; **A. van Hylckama-Vlieg**: Conceptualization, Supervision, Writing – Reviewing and Editing; **N. A. Aziz**: Conceptualization, Methodology, Writing – Review and Editing; **R. Li-Gao:** Writing – Review and Editing; **R. de Mutsert**: Project Administration, Resources, Funding Acquisition, Writing – Review and Editing; **F.R. Rosendaal.**: Study design, Funding acquisition, Conceptualization; **R. Noordam** and **D. van Heemst**: Funding acquisition, Writing – Review and Editing; **D.O. Mook-Kanamori:** Conceptualization, Methodology, Resources, Writing – Reviewing and Editing, Funding Acquisition, Supervision; **K.W. van Dijk**: Conceptualization, Supervision, Writing – Reviewing and Editing; **M.M.B. Breteler:** Conceptualization, Methodology, Resources, Writing – Reviewing and Editing, Data Curation, Funding Acquisition, Supervision.

All authors read and approved the final manuscript

### Data availability

Due to the privacy of the participants of the NEO study and legal reasons, we cannot publicly deposit the data. Furthermore, NEO study participants did not sign informed consent to make their data publicly available. Data can be made available upon request to interested qualified researchers. Data requests should be sent to the NEO Executive Board which can be contacted via https://www.lumc.nl/org/neo-studie/contact/.

Rhineland Study data used for this manuscript are not publicly available due to data protection regulations. Access to data can be provided to scientists in accordance with the Rhineland Study’s Data Use and Access Policy. Requests for additional information and/or access to the datasets can be send to.

All authors had full access to their respective study data and take responsibility for the integrity of the data and the accuracy of the analysis.

### Ethics approval

The NEO study was approved by the medical ethical committee of the Leiden University Medical Centre (LUMC) and all participants gave their written informed consent.

The ethics committee of the medical faculty of the University of Bonn approved the undertaking of the Rhineland Study and it was carried out according to the recommendations of the International Council for Harmonisation Good Clinical Practice standards.

### Consent to participate

Written informed consent was acquired from all participants per the Declaration of Helsinki in both the NEO and Rhineland Study.

## Supplementary Figures

**Supplementary Figure 1:**
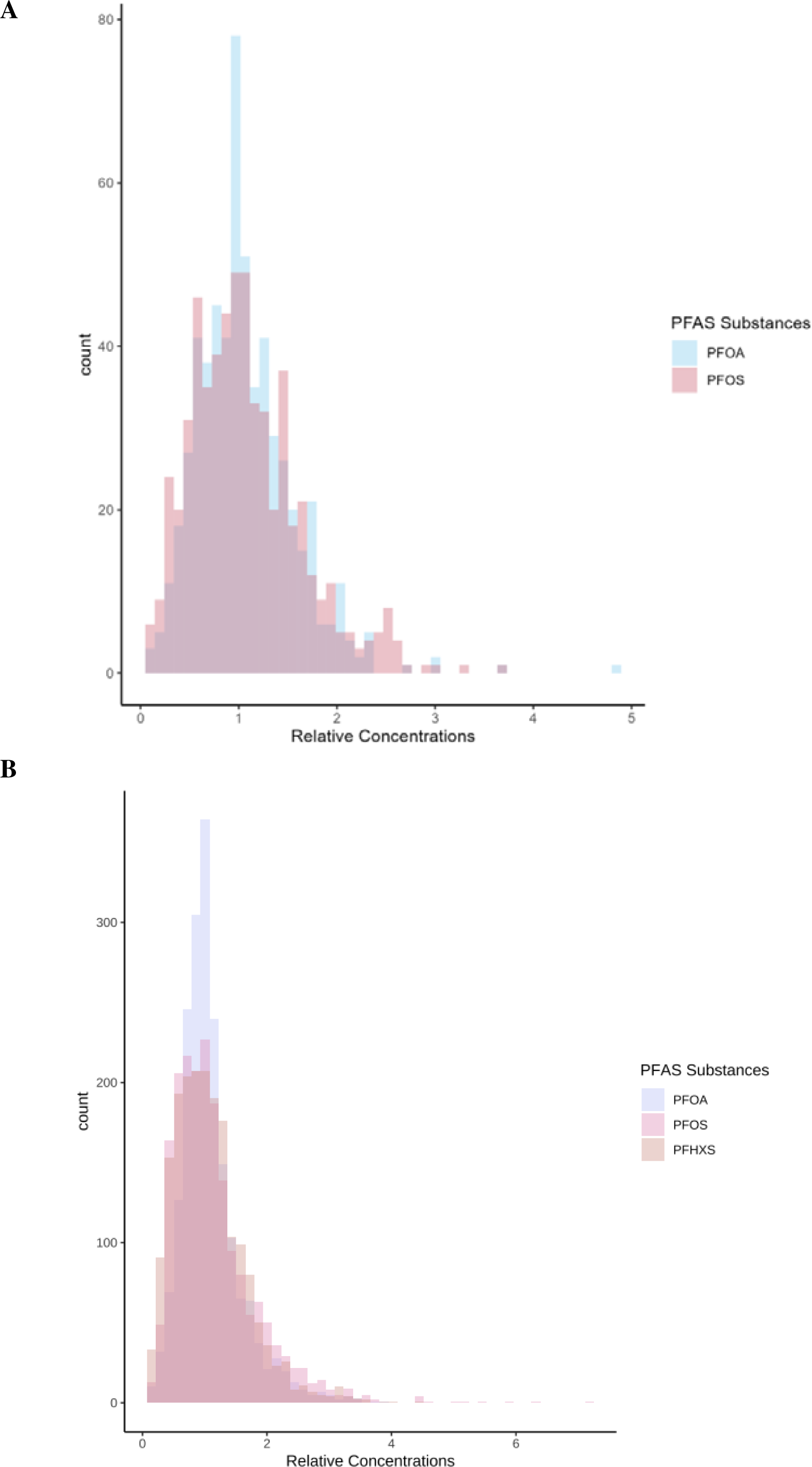
Distribution of the PFAS levels in NEO (A) and the Rhineland Study (B), where outliers (>5 standard deviations) are truncated for PFOA (n= 9), PFOS levels (n= 5) and PFHxS (n= 4) in the Rhineland Study.

